# Machine Learning and Prediction of All-Cause Mortality among Chinese Older Adults

**DOI:** 10.1101/2021.04.21.21255843

**Authors:** Xurui Jin, Yiyang Sun, Tinglong Zhu, Yu Leng, Shuyi Guan, Kehan Zhang, Kangshuo Li, Zhangming Niu, Chenkai Wu, Yi Zeng, Yao Yao, Lijing L. Yan

**Author notes:** Corresponding authors: Professor Lijing L. Yan., Global Health Research Center, Duke Kunshan University, Jiangsu Province, China., Dr. Xurui Jin, Global Health Research Center, Duke Kunshan University, Jiangsu Province, China.

## Abstract

**Background and aim:** Mortality risk stratification was vital for targeted intervention. This study aimed at building the prediction model of all-cause mortality among Chinese dwelling elderly with different methods including regression models and machine learning models and to compare the performance of machine learning models with regression model on predicting mortality. Additionally, this study also aimed at ranking the predictors of mortality within different models and comparing the predictive value of different groups of predictors using the model with best performance.

**Method:** We used data from the 2008 and 2011 waves of Chinese Longitudinal Healthy Longevity Survey (CLHLS). The follow-up of CLHLS was conducted every 2-3 years till 2018. The analysis sample included 2,448 participants. We used totally 117 predictors to build the prediction model for survival including 61 questionnaire, 41 biomarker and 15 genetics predictors. Four models were built (XG-Boost, random survival forest [RSF], Cox regression with all variables and Cox-backward). We used C-index and integrated Brier score to evaluate the performance of those models.

**Results:** The XG-Boost model and RSF model shows slightly better predictive performance than Cox models and Cox-backward models based on the C-index and integrated Brier score in predicting surviving. Age, activity of daily living and Mini-Mental State Examination score were identified as the top 3 predictors in the XG-Boost and RSF models. Biomarker and questionnaire predictors have a similar predictive value, while genetic predictors have no addictive predictive value when combined with questionnaire or biomarker predictors.

**Conclusion:** In this work, it is shown that machine learning techniques can be a useful tool for both prediction and its performance sightly outperformed the regression model in predicting survival.

## Introduction

Currently, aging is a global phenomenon, and in particular, the proportion of older individuals in low and middle-income countries (LMIC) is growing in an accelerated pace due to the increasing average life span. Among LMICs, China shows the fastest rate of aging, and the related health care cost has been growing rapidly. According to the UN statistics database, China’s dependency ratio for retirees could rise as high as 44% by 2050 which would then be the highest around the world [1]. As the population is aging quickly, it is important to have adequate identification and risk stratification for individuals with reduced life expectancy. Adequate risk stratification of mortality can lead to a more precise intervention and more targeted care. It also has important clinical relevance as an accurate mortality risk assessment tool might improve the accuracy of the prognostic assumptions which in turn can influence clinical decisions [2, 3].

Most of the current estimation methods available for mortality prediction are based on a single independent factor including blood pressure [4], body weight [5], walking speed [6], self-reported health [7] and frailty [8]. There are some indices consisting of several predictors for short-term mortality, but they have mainly been developed for and assessed in older individuals or in high-risk populations [3]. To be more specific, a large number of clinical conditions were assessed their relevance in the prediction of 1-year mortality and they were summarized into the Charlson Comorbidity Index (CCI) [9], which has been validated in large populations and widely used to predict mortality among hospitalized patients. However, since more data, including biomarkers and genetic assessments, became available, a systematic approach has become a trend in medical and public health studies. However, it is still unclear whether only using several or only a selected group of predictors for prediction has limitations in providing accurate mortality risk stratification. Additionally, an inaccurate mortality risk stratification may lead to an untargeted mortality prevention strategy [2].

Following the fast-paced development of biomedical technology, more and more high dimensional data became available in clinical and public health studies [10]. These data include genetics, metabolomics, and proteomics. Additionally, the number of predictors in epidemiological studies were also increasing. Normally, hundreds of predictors are available for developing mortality prediction models in epidemiological studies or electronic medical record data. The core of traditional statistical methods is hypothesis testing and this process is user-driven which indicated that the researcher would have to specify dependent variables, regression family and link and interaction type. Therefore, user intervention may influence the results of those models. Another methodological concern was that the traditional statistical techniques such as regression models are often limited by the correlation between variables, nonlinearity of variables, and the possibility of overfitting [11, 12]. The newly developed machine learning method has provided the possibility the address those challenges. With machine learning techniques, the hypothesis is that the associations between various predictors and the outcome were a pattern [13]. Thus, it is a hypothesis-free method. Machine learning models can analyze all predictor variables in a way that prevents overlooking potentially important predictor variables even if it was unexpected. Accordingly, it is reasonable that machine learning method could be an effective method in identifying the best predictors of outcomes from a large number of predictors.

Most of the prediction models are based on regression model and the researcher need to manually input the predefined interactions, such as the interaction between vitamin D and albumin on mortality I have demonstrated [14]. Missing those complex interactions in the regression model may result in an inaccurate prediction of outcomes. In the prediction model developed by machine learning methods, the model can automatically identify those interactive relationship from the data and it is unnecessary to specify interactions [11, 12]. However, it is still widely debated whether the performance of machine learning model was better than that of regression model and recently it was a very hot issue to demonstrate the usefulness of these methods.

Recently, a systematic review that included 71 from 927 searched studies suggested that some common-used machine learning methods were classification trees, random forests, artificial neural networks, and support vector machines [12]. It was found that the difference in logit between logistics regression and machine learning was 0.00 (95% CI: −0.18, 0.18). Moreover, a meta-analysis published in Nov. 2020 suggested that machine learning models provide better discrimination in mortality prediction after cardiac surgery [11]. However, among those studies included in those two reviews, the number of predictors and the sample has a very large variation. Therefore, more studies are warranted to compare those two methods in different cohorts and different populations.

The overarching goal of this study is to provide mortality risk stratification tools for the Chinese older adults. Specifically, there are three major research objectives and their sub-objectives of this study:

1. Using the data from CLHLS with maximal number of predictors to build the survival prediction model for Chinese older adults using different methods including regression models (Cox and Cox-backward) and machine learning models (random survival forest [RSF] and XG-boost) and to compare the performances of machine learning models with regression models in predicting survival.
2. To rank the predictors of mortality within different survival models.
3. To compare the predictive values of different groups of predictors using the best performing model

## Method

### Study population

The present study uses data from the Chinese Longitudinal Healthy Longevity Survey (CLHLS). CLHLS is a longitudinal study since 1998 with follow-up surveys every 2-3 years till 2018. The CLHLS surveys were conducted in randomly selected counties and cities in China, which accounted for half of the counties and cities in 23 out of 31 provinces covering over 85% of China’s population. Based on gender and place of residence (ie, living in the same street, village, city or county) for a given centenarian, randomly selected octogenarians and nonagenarians were also sampled. This matched recruitment procedure resulted in an oversampling of the oldest old and older men. In the CLHLS, a weight of age-sex urban/rural residence in the sample with the distribution of the total population in the sampled 22 provinces was employed to reflect the unique sampling design. More details of this survey have been published elsewhere [15]. In this study, we derived data from the CLHLS biomarker study. The CLHLS biomarker study was conducted in eight longevity areas [16]. This study covers similar domains that CLHLS has investigated and shared the sampling strategy as CLHLS. Participants with age less than 65 years old (n=51) and with missing values in higher than 30% of the predictors were excluded (n=33) and my analyses included 2,448 elderly aged 65 years or over who had both phenotypic, biomarker, and genotypic measurements.

The Ethics approval of CLHLS biomarker study was obtained from the Research Ethics Committees of Peking University and Duke University. All participants or their legal representatives signed written consent forms in the baseline and follow-up surveys.

### Predictors and imputation techniques

After excluding those variables with missing value higher than 30%, 117 predictors were included in this study including demographic variables, lifestyle (smoking, drinking, diet and physical activities), health indicators (cognitive function, activity of daily living and leisure activities), comorbidities, biomarkers and genetic information.

The questionnaire data were collected through in-home interviews by trained interviewers who are local staff members from the county-level network system of the National Bureau of Statistics of China. All interviewers have received 12+ years of schooling, and most have earned a college degree. Each interviewer was accompanied by a local doctor, a nurse, or a medical college student so that some health check-ups could be performed. In the physical examination, body weight and height were measured by trained medical staff using a standardized protocol. Totally 61 questionnaire predictors were included in the analysis.

The CLHLS collected blood and urine sample since 2008. During the investigations, blood samples were centrifuged within 1 hour after collection and heparin anticoagulant blood samples were centrifuged at 3000 rpm for 10 min at 18°C–25°C. Then blood and urine samples are immediately stored at −80°C in the local Center of Disease Control and Prevention (CDC). And then the sample were transported at −20°C with transport cases provided by CCDC by specially assigned persons to designated testing units. A variety of blood and urine biomarker were measured and all laboratory analyses are conducted by the central clinical laboratory at Capital Medical University in Beijing. The protocol of biomarker measurement was published elsewhere. In this study, totally 41 biomarker predictors were included. [16]

The genotyping was performed by a customized chip targeting about 287,898 candidate SNPs associated with longevity, chronic disease or health indicators based on multiple studies. There were no familial/kinship relations among the participants within and across different waves. Beijing Genomics Institute (BGI) performed the genotyping, and the BGI genotyping quality control procedures of the genetics study have been published elsewhere. [17] In this study, totally 15 SNPs were selected by previous meta-Genome wide association studies (GWAS) focusing on longevity and our previous GWAS study using the same data [17-20]. All the predictors are listed in Table 1.

**Table 1.**
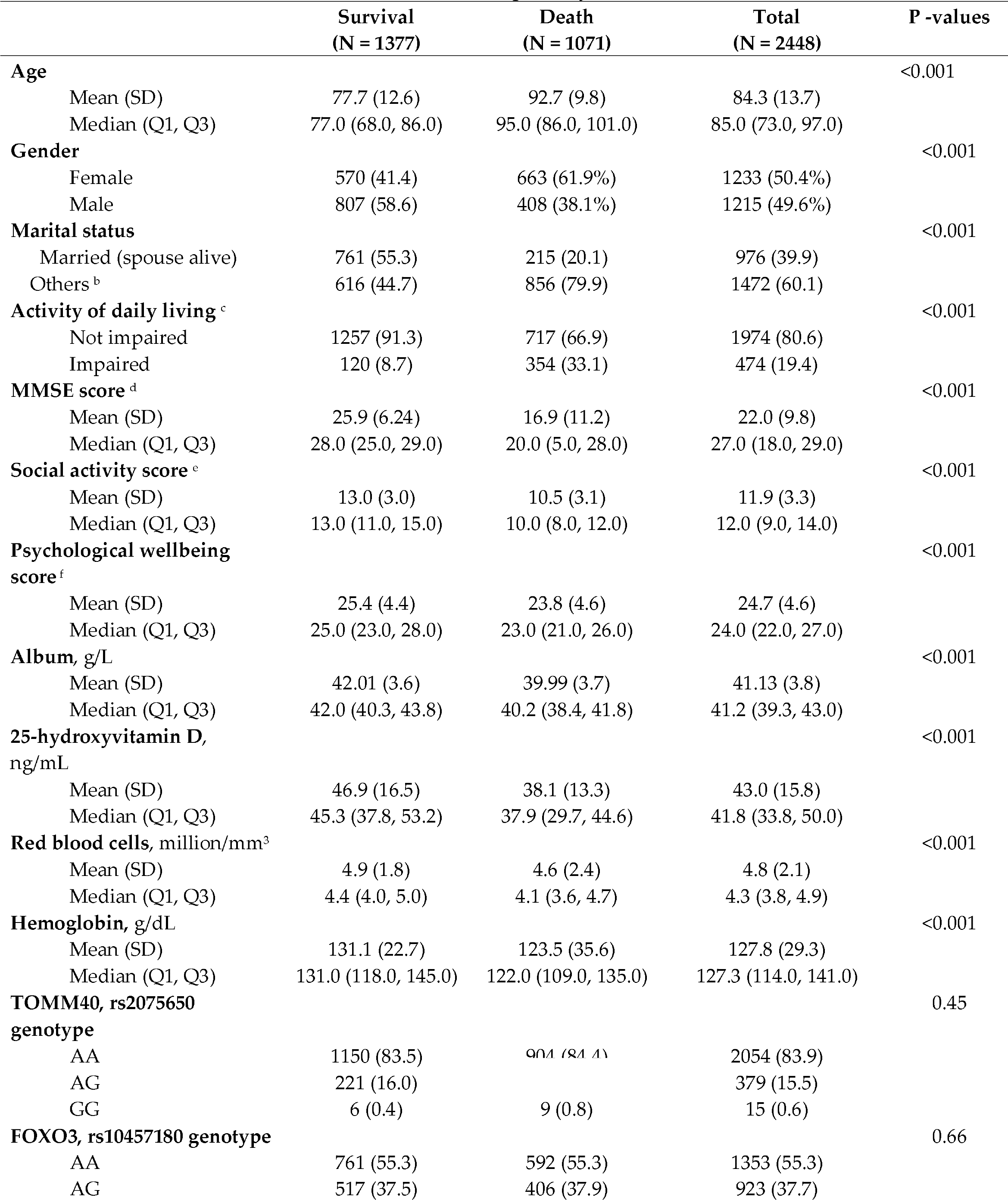
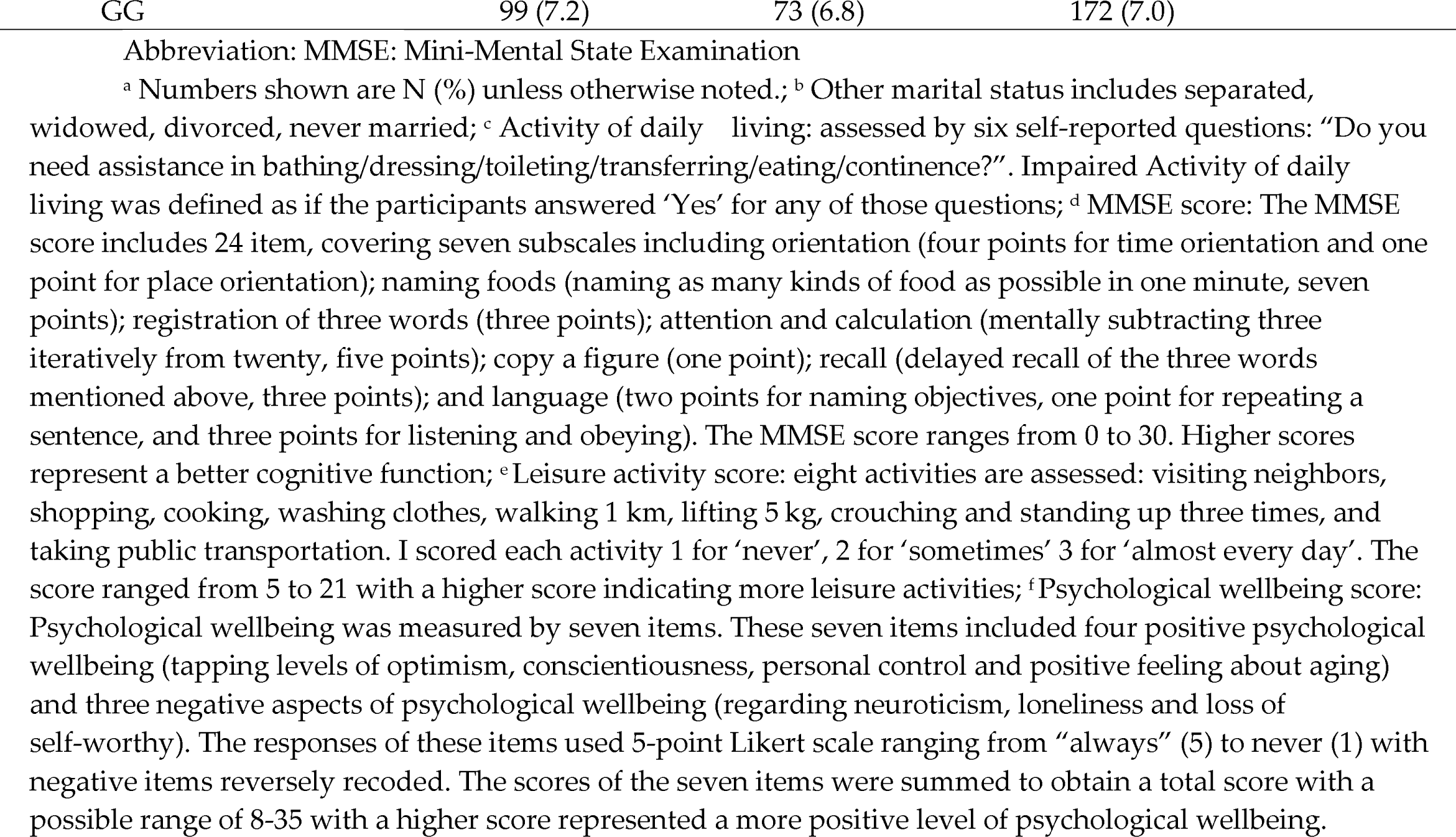
Baseline Characteristics ^a^ of the CLHLS Participants by Survival Status.

Overall, there are 4.7% missing data. We use the missForest algorithm to impute those missing values [21]. This method is a nonparametric imputation method that builds a random forest model for each variable and it was demonstrated that this method outperformed many imputation methods especially in data settings where complex interactions and non-linear relations are existing.

### Data on mortality

Vital status and date of death were collected from officially issued death certificates when available or otherwise from the next-of-kin or local residential committees who were familiar with the decedents. Duration of follow-up was calculated by the time interval between the first interview date and date at death. Survivors at the last wave (2018) were censored at the time of the last survey.

### Statistical analysis

Totally four models for predicting survival were built: 1. The RSF model, 2. the XG-boost model, 4. the Cox model with all predictors, 4. the COX-backward model. For the analysis using data from CLHLS, I build the XG-boost model to predict the two years’ mortality.

In order to train and validate the models and optimize the machine learning models, I used 10-fold internal cross validation. The training data was randomly divided into ten folds, and each time, nine folds of data were included in the training model and the rest one fold of data was used as the testing test. This process was repeated ten times for all combinations of folds. I used the grid search to determine the hyper-parameters of machine learning model.

To evaluate the performance of those models, I use Harrell’s concordance index (C-index) and integrated brier score (IBS) for the survival model. Higher C-index and lower IBS/BS indicate better discrimination and calibration performance.

#### Cox proportional hazard regression models and Cox-backward models

In this survival analysis, the focus is on the period till the occurrence of mortality. The Cox proportional hazards model is usually used to estimate the hazard ratio of the interested factors on the outcome. I build two Cox models. One is the Cox with all predictor and the other is the Cox model with a backward elimination model (Cox-backward). The Cox-backward model was widely used for variable selection. I used R (version 3.6.1) to build the Cox model.

#### XG-boost

In the survival analysis and the analysis of predicting two year’s mortality, I built the extreme gradient boosting (XG-Boost), an ensemble machine learning method based on decision trees, to establish the prediction model for mortality with all the predictors. Gradient boosting is a machine learning model that involves combinations of prediction models into a strong model. The XG-Boost approximates the value of the loss function with the second-order Taylor series and reduces the probability of overfitting by regularization.

Some hyper-parameters parameters of XG-Boost model was defined as following: 1) number of trees: 400, learning rate: 0.005, 2) minimal loss to expand on a leaf node: 0; 3) maximum tree depth: 4, 4) subsample proportion: 1. Additionally, the XG-Boost model can provide the estimations of feature importance from the trained model. In this study, I used F scores in XG-Boost model to evaluate the feature importance which is the sum of Gini index among the corresponding splits in a tree and further averaged among all the trees. Python 3.7 was used to build this model.

#### RSF

In the survival analysis, I build the RSF model with all predictors. RSF designed for time-to-event data such as survival as a transformed RF. Some hyper-parameters parameters of RSF model were defined as following: 1) number of trees: 1000, number of random split points used to split a node: 5, 2) mtry value: 12; 3) node size: 50, 4) block size: 5. I applied variable importance (VIMP) to ranking the predictors. The absolute value of VIMP indicates the impact of this predictor on the overall performance. The positive VIMP value indicates the predictor improves predictive performance and negative value indicates that the predictor has negative effect on the performance of the prediction model. I used “randomForestSRC” and “cph” in R (version 3.6.1) to perform those analysis.

## Results

### Baseline characteristics

The median follow-up period was 3.6 years (range: 0.2-9.9 years). Totally 43.8% (n=1,071) of all participants died during the follow up. Table 1 presents the part of baseline characteristics by survival status at the last follow up. The mean age was 84.3 years (SD: 13.7). Participants who were survival are more likely to be younger, male, without impaired activity of daily living, married, with higher MMSE score, social activity score and psychological wellbeing score, with higher levels of album, 25-hydroxyvitamin D, red blood cells and hemoglobin (Ps<0.05).

### Comparisons between models

Two Cox models were built: a) a model with all 117 predictors and a Cox model with backward selection. Furthermore, two machine learning models were built: a) a RSF model, b) XG-Boost model.

**Table 3** presented the performance of the four models on test data. Regarding the IBS, the XG-Boost models have the lowest (IBS = 0.120) followed by the RSF (IBS = 0.128). Cox models have sightly higher IBS (COX with all variables: IBS = 0.131; Cox-backward: 0.129). In terms of C-index, the XG-Boost model has the highest C-index (C-index: 0.80), and the Cox models with all variables has slightly worse performance (Cox model with all variables: C-index: 0.77; Cox backward model: C-index: 0.78). C-index for Cox backward and RSF models are nearly the same (C-index: 0.78).

**Figure 1** shows the average prediction Brier error over time for those four models (XG-Boost, RSF, Cox with all variables and Cox-backward). Only small differences can be observed between Cox and Cox-backward models. The XG-Boost model achieved better performance than the other models, but only sightly better than the RSF model.

**Figure 1:**
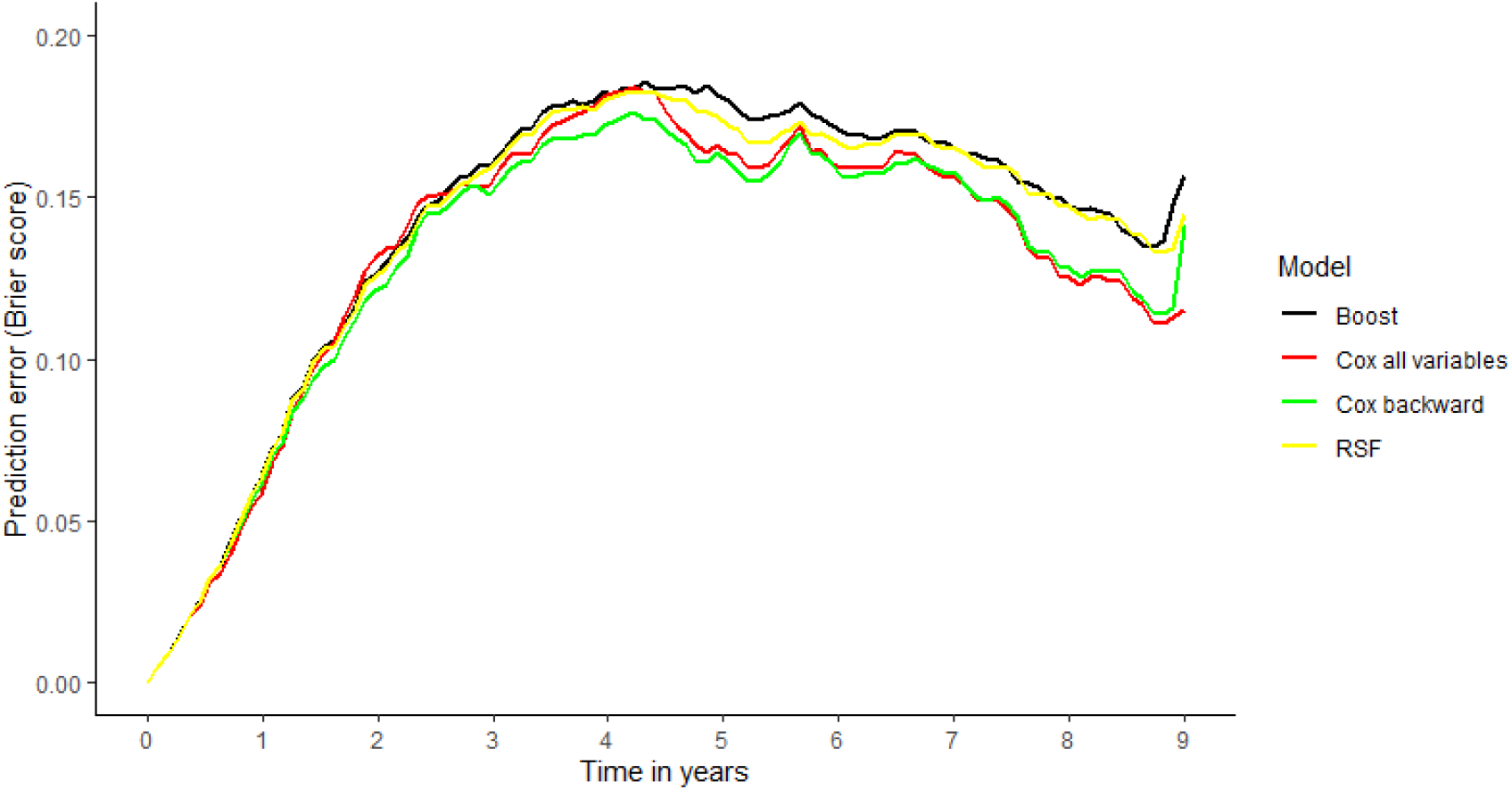
Prediction Error Curves for all Models.

**Figure 2:**
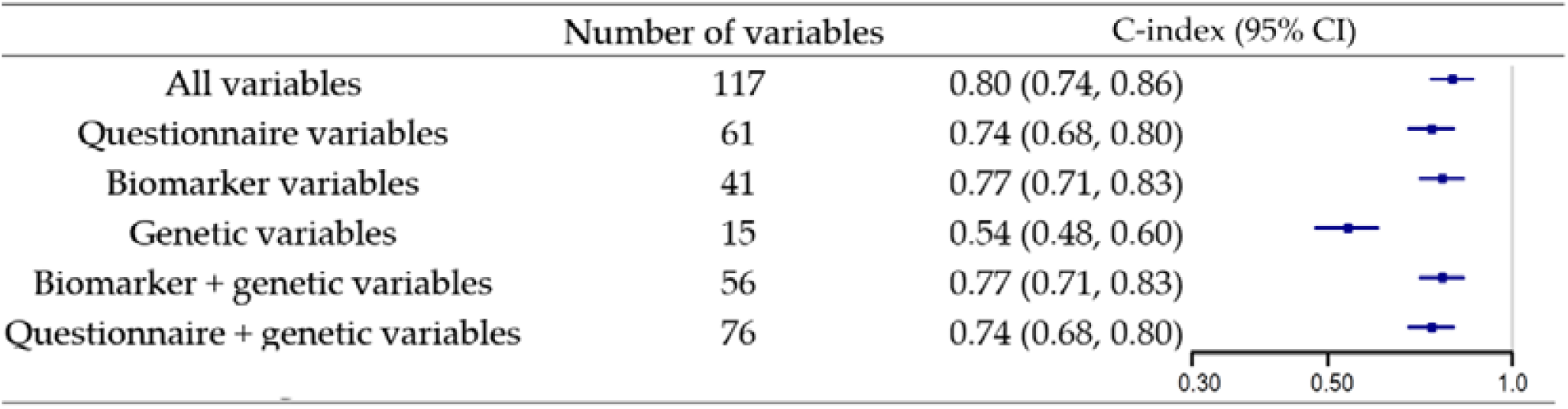
The C-index and 95% Confidence Interval of XG-Boost model with different groups of variables.

### Predictor ranking

Hazard ratios of the 5 most predictive variables for the Cox models with all predictors are shown in **Table 2** which was selected by the z-score values. The strongest predictor is chronological age. One-year increase in age increased a 5% higher mortality risk. The other most predictive variables are mean corpuscular hemoglobin concentration (One-unit increase: HR: 1.005, 95% CI: 1.002, 1.008), gender (male: 1.66, 95% CI: 1.23, 2.24), activity of daily living (impaired: HR: 1.40, 95% CI: 1.11, 1.77), urine microalbumin (one-unit increase: HR: 1.002, 95% CI: 1.000, 1.003). In the Cox backward model, after the backward selection, there are only five variables left which are urine microalbumin (HR: 1.003, 95% CI: 1.002, 1.004), MMSE score (HR: 0.98, 95% CI: 0.97, 0.99), age, (HR: 1.06, 95% CI: 1.05, 1.07), albumin (HR: 0.93, 95% CI: 0.90, 0.95), 25-hydroxyvitamin D (HR: 0.98, 95% CI: 0.97, 0.98). The predictor with highest predictive value was urine microalbumin.

**Table 2.** A List of the Predictors Used for Prediction.

|  |
| --- |
| **Questionnaire: basic demographics and lifestyle** |
| Age, gender, ethnicity, co-residence, years of schooling, marital status, residence, occupation before retirement, smoking status, drinking status, physical activity, consumption of fruit, vegetable, meat, fish, egg, bean, salt-preserved vegetables, sugar, tea, garlic, height |
| **Questionnaire: disease history and health indicators** |
| Times of suffering from serious illness in past two years, suffering from hypertension, diabetes, heart disease, stroke, bronchitis, emphysema, pneumonia, asthma, tuberculosis, cataract, glaucoma, cancer, ulcer, Parkinson's disease, bedsore, arthritis, systolic and diastolic blood pressure, BMI, self-reported health, self-reported life satisfactory, activity of daily living, social and leisure activities, MMSE score, psychological well-being |
| **Biomarkers** |
| White blood cell count, red blood cell count, hemoglobin, erythrocyte hematocrit, erythrocyte mean corpuscular volume, erythrocyte mean corpuscular hemoglobin, erythrocyte mean corpuscular hemoglobin concentration, platelet count, plateletocrit, mean platelet volume, lymphocyte count, percentage of lymphocytes, platelet distribution width, white blood cell leukocytes, nitrates, urobilinogen, bilirubin, ketones, glucose, Vitamin C, high-density lipoprotein cholesterol, low-density lipoprotein cholesterol, urea acid, plasma creatine, urea nitrogen, triglyceride, total cholesterol, high sensitive c-reactive protein, malondialdehyde, superoxide dismutase activity, albumin, creatine, glycolated plasma protein, superoxide dismutase, 25-OH-D3, vitamin B12, urine microalbumin, urine creatinine, urine microalbumin/ urine creatinine ratio |
| **Genetic** |
| HLA-DQA1/DRB1: rs34831921, TOMM40: rs2075650, APOC1: rs4420638, NECTIN2: rs6857, APOE: rs769449, BEND4: rs1487614, EPHA6: rs10934524, ZFYVE28: rs57681851, ASIC2: rs7213812, FAM13A: rs2609284, LIMCH1: rs10007810, FOXO3: rs10457180, OR2W3: rs10888267, LINC02227: rs2149954, CSRNP3: rs6432832 |
| Abbreviation: MMSE: Mini-Mental State Examination |

**Table 3** presented the top-10 predictors in the RSF and XG-Boost models. The importance of predictors is assessed by F score in the XG-boost models and VIMP in RSF. The strongest predictor is age in the two models. In those top predictors, age, MMSE score, activity of daily living, social activity score, 25-hydroxyvitamin D, albumin, marital status and hemoglobin were both identified in those two models. “Red blood cell” was identified as high predictive value in RSF model (VIMP: 0.0023) and “self-reported health” was identified by XG-Boost model (F score: 0.0018).

**Table 3:** Integrated Brier Score (IBS) and C-index on the Test Data.

| CLHLS | IBS | C-index |
| --- | --- | --- |
| **Cox all variables** | 0.131 | 0.77 |
| **Cox backward** | 0.129 | 0.78 |
| **RSF** | 0.128 | 0.78 |
| **XG-boost** | 0.120 | 0.80 |

**Table 4:** Hazard ratios along with their 95% confidence intervals for the 12 most influential variables for the Cox models.

| Variable name | Cox all variables,<br>HR (95% CI) | Variable name | Cox backward,<br>HR (95% CI) |
| --- | --- | --- | --- |
| One-year increase in age | 1.05 (1.04, 1.06) | One-unit increase in<br>urine microalbumin | 1.003 (1.002, 1.004) |
| One-unit increase in mean corpuscular<br>hemoglobin concentration | 1.005 (1.002, 1.008) | One point increase in<br>MMSE score | 0.98 (0.97, 0.99) |
| Gender, male | 1.66 (1.23, 2.24) | One-year increase in<br>age | 1.06 (1.05, 1.07) |
| Activity of daily living, impaired | 1.40 (1.11, 1.77) | Albumin | 0.93 (0.90, 0.95) |
| One-unit increase in urine microalbumin | 1.002 (1.000, 1.003) | 25-hydroxyvitamin D | 0.98 (0.97, 0.98) |

Variables are presented in decreasing order according to the absolute z-score values for the Cox model with all variable

**Table 5:** The 10 most prognostic factors for the XG-Boost and for the Random Survival Forest.

| Random Survival Forest |  | XG-Boost |  |
| --- | --- | --- | --- |
| Variable | VIMP | Variable | F score |
| Age | 0.0389 | Age | 0.068 |
| MMSE score | 0.0150 | Activity of daily living | 0.024 |
| Activity of daily living | 0.0137 | MMSE score | 0.023 |
| Social activity score | 0.0095 | Social activity score | 0.021 |
| 25-hydroxyvitamin D | 0.0055 | 25-hydroxyvitamin D | 0.0068 |
| Albumin | 0.0032 | Marital status | 0.0048 |
| Marital status | 0.0030 | Psychological wellbeing score | 0.0029 |
| Red blood cells | 0.0023 | Self-reported health | 0.0018 |

Abbreviations: VIMP: variable importance

### Comparisons between groups of predictors

The predictors were grouped as “questionnaire variables”, “biomarker variables” and “genetic variables” and I put different groups of predictors in the XG-Boost model which has the best predictive performance. When solely add “questionnaire variables”, “biomarker variables” or “genetic variables” into the model, the C-index for each XG-Boost model was 0.74 (95% CI: 0.68, 0.80), 0.77 (0.71, 0.83) and 0.54 (95% CI: 0.48, 0.60) respectively. After adding the genetic variables into the XG-Boost model with questionnaire or biomarker variables, the predictive value did not improve and the C-indexes before and after adding the genetic variables was nearly the same.

## Discussion

In this large, prospective cohort study, with the data form CLHLS biomarker cohort (N=2,448, 117 predictors), I did an extensive analysis of mortality prediction model which included over one hundred predictors with all-cause mortality followed up to ten years. I have ranked the predictive value of those predictors from questionnaire, biomarker and genetic assessment.

Several key messages can be concluded from this study. First, the performance of machine learning model was sightly better than the regression model in predicting survival. Second, predictors that can simply be obtained by interview without blood testing can predict all-cause mortality with a high predictive value among the older population. Age, activity of daily living and MMSE score were the strongest predictors. Third, generally, the predictive value of biomarker predictors was highest, and the predictive value of questionnaire predictors was comparable, however the predictive value of genetic predictors was low, and it did not increase the performance of prediction model when combined with other groups of predictors. In this report, I have presented only part of my findings, and a detailed version of my results are available in an open access database where the detailed predictive value for each variable in each model was available to generate new research hypotheses for other researchers.

In this study regression and machine learning models were applied for predicting all-cause mortality among Chinese community-dwellings within two cohorts. Generally, I found that machine learning models outperformed the regression model in predicting mortality in terms of C-index and IBS, but the difference was quite small. The results indicated that machine learning methods are well suited for meaningful risk prediction in large-scale epidemiological studies. Theoretically, compared with regression models, machine learning methods can avoid the problem of overfitting and non-convergence, and also considering the non-linearities. However, it is still controversial that whether machine learning could improve the accuracy of mortality risk stratification. In some study with relatively small sample size and limited number of predictors, the regression models have a comparable performance as the machine learning models. In one study including 1701 men with a follow up of 3 years and eight predictor available [22], it found that in predicting survival, machine learning models did not have a better performance as the regression models (C-static: Cox-regression: 0.78; survival tree: 0.71; binary tree: 0.66; logistics regression: 0.72) In another study including 603 patients from the hospital with ST elevation myocardial infarction, using 10-fold cross validation, the logistics regression achieves the highest C-statistics of 0.82 and outperformed decision tree, naive Bayes classifier, artificial neural network and Bayesian network classifier. However, in some studies with larger simple size and larger numbers of predictor, machine learning models outperformed the regression model. For instance, a study including 6,520 patient with 66 predictors compared the performance of logistic regression model and different machine learning models on predicting the mortality in-hospital after elective cardiac surgery [23]. Four different machine learning models have been evaluated with the regression models: gradient boosting machine, random forest, support vector machine and naive bayes. The area under the ROC curve for the machine learning model (C-index = 0.80) was significantly higher than the logistic regression model (C-index = 0.742). Similar result was found in another study with a large size of simple and predictors, a study derived data from the Multi-Ethnic Study of Atherosclerosis (MESA) including 6814 participants aged 45 to 84 years [24]. Seven-hundred thirty-five variables from imaging and noninvasive tests, questionnaires, and biomarker panels were used to build the RSF and Cox model. In predicting all-cause mortality, the RSF model has a higher C-index (0.86) compared with that of the Cox regression models (AIC-Cox with forward selection: 0.78; LASSO-Cox: 0.80). To conclude, it is plausible that when the sample size was small and the number of predictors, the performance of machine learning model seems comparable with the regression model, while when the sample size was big enough and the number of predictors was larger enough, the performance of machine learning model will outperform the regression model. Combined with previous evidence, our analysis with the sample of over two thousand elderly with over one hundred predictors also partly demonstrated this hypothesis. However, further methodology studies were warranted to investigate in what situation the machine learning models would outperform the regression model. Additionally, the methodology development of the interpretative machine learning method is also needed to understand how those interactive relationships influent the predictive performance of the models. As we move into the age of precision medicine, understanding the use of phenotypic data and methods to analyze already acquired information is of paramount importance.

Previous studies were mainly focus on the impact of signal predictors and could not have a comparison among those predictors, while our analysis provided the information about the relative importance of each variable as predictor of all-cause mortality. Several previous studies on mortality have ranked the predictors from across domains. In a study using data from UK biobank, 655 predictors were evaluated on their predictive value of five-year mortality in nearly 500,000 adults aged over 50 years old [25]. Those predictors included blood biomarkers, disease histories, socio-demographics, early life health factors and family history, psychosocial factors, and healthy lifestyle. Among those predictors, self-reported health was the strongest predictors of all-cause mortality. In another study derived data from Heath and Retirement Study [26], totally 57 predictors of adverse socioeconomic and psychosocial experiences during childhood, socioeconomic conditions, health behaviors, social connections, psychological characteristics, and adverse experiences during adulthood was evaluated. Smoking was the strongest predictor of mortality among those 13,611 American aged from 52 to 104 years old. Of note, in those two studies examined the predictive value of a comprehensive groups of predictors, age was regarded as covariate. In another study using machine learning methods the most powerful predictor for all-cause mortality was the chronological age. Additionally, in that study, some novel biomarkers were identified as tissue necrosis factor-α soluble receptor and interleukin-2 soluble receptor [24]. In my study, I have also identified some other predictors need to be addressed such as activity of daily living as a measurement of activity of daily living, blood pressure and MMSE score as a measurement of cognitive function which was previously underestimated in the mortality risk prediction models. This results were corresponding with the findings of UK biobank study which indicate that some general health indicator may be the most potent mortality predictor [25]. To note, the importance of cognitive health and blood pressure control were addressed in my study. In China, A large number of elderly people are expected to suffer varying degrees of cognitive impairment and hypertension in the future.[27] My findings along with evidence on the lack of adequate care for cognitive impairment and blood pressure control in China indicate that there is an urgent need for China’s health care system and government to improve provision and quality of these services. My study also suggested that some biomarkers such as red blood cells, hemoglobin and urine microalbumin which was rarely studied also need to be addressed in the future mortality risk stratification studies. In terms of those finding, machine learning enabled the researcher to discover new relationships without prior without prior assumptions. Identifying effective mortality risk predictor may be of benefit for effective screening strategies and suggest specific targets for risk reduction. Additionally, although I have identified some predictors which seems not actionable on the personal level such as cognitive function and social and leisure activity. But this should not be interpreted as an impediment to improvement of health behaviors. Previous studies have demonstrated that healthier modifiable behaviors such as quit smoking, higher level of physical activities and healthier diet are beneficial and further reduce the risk of mortality.

Another finding of my analysis was that the predictive value of questionnaire and biomarker predictors was comparable, and the predictive value of genetic predictor was low. Additionally, adding genetic variables into the prediction model with biomarker or questionnaire predictors did not substantially improve its performance. Our results were corresponding with another study which also suggested that the predictive value of genetic predictors was limited. The study included 5,974 participants from the Rotterdam Study [28], followed for a median of 15.1 years, and it has demonstrated that specific genetic factors were independently associated with mortality, jointly they contributed little to mortality prediction (C-index = 0.56). Combined with those previous studies, it is possible that common SNPs only have very limited predictive power of mortality or longevity, when comparing with those traditional predictors. Considering its limited predictive power, it may be not necessary to perform the genetic assessment when evaluating the mortality risk. Although my results suggested that it may be unnecessary or invalid to assess the individual’s genetic background in mortality risk prediction, it is still need to be validated for some specific disease risk prediction such as the APOE gene in predicting the risk of dementia and BCL2 gene in predicting breast cancer. Additionally, it may be possible that some epigenetic biomarker such as the methylation in genomes may improve the predictive performance of prediction models, however the cost of such measurement was much higher and the cost-effectiveness needs to be considered.

This is the first study where machine learning models are applied to data from Chinese dwelling elderly in predicting all-cause mortality where a comparison with the traditional Cox model was also performed. My study has some implications for at the individual, clinical or policy making level especially for those at the LMICs with a rapid pace of aging.

At the individual level, it can be applied to address the dwelling’s self-awareness of the health. For example, taking more health behaviors such as quit smoking and alcohol drinking to prevent cognitive impairment and control the blood pressure. At the clinical and community level, the physician or health worker might use the model to identify individuals who were at high risk of mortality. To note, currently, frailty, as a measure consisting nearly one hundred variables, was gradually used in the real-world settings to perform the mortality risk stratification. However, it might be possible that my model which would be more convenient, as the development of medical informatics, could be applied to evaluate the mortality risk in different settings and may outperformed the frailty measurement. With such mortality risk stratification, targeted specific interventions or treatment can be implemented to those individuals and further improve the quality of healthcare. Finally, policy maker can use this information to allocate more medical or public health recourse to decrease the burden of specific risk factors.

My study has several limitations. Firstly, the sample size is limited of this study, but, it has included a wide range of predictors. It is difficult to find another cohort with such big numbers of predictors. Secondly, the accuracy of the predictor. I used many self-reported predictors and those predictors are always subject to misclassification bias. However, the data of CLHLS has a good overall quality and its reliability and validity were validated. Thirdly, the genetic information was selected from the GWAS studies, which may not fully represent the genetic risk of mortality, however, I extracted the genetic information according to the published GWAS study using the same data, which may reduce the under estimation of the genetic mortality risk. Fourth, some biomarker was not available in my data such as the Interleukin 6 or Tumour Necrosis Factor alpha which represented level of inflammation, however, in our data, there are other biomarker measured the inflammation level such as albumin and high-sensitivity C-reactive protein. Fifthly, I use the internal validation method which is the training and test data sets were both drawn from the CLHLS study population. An external validation with data from other cohorts was needed.

## Conclusions

In this population-based study, I built the prediction models with machine learning models and regression model and I found that machine learning model sightly outperformed the regression model in predicting all-cause mortality. Age, MMSE score and ADL were three of the most important predictors. Additionally, I have compared the predictive value of different groups of predictors and found that the predictive value of genetic predictors was much lower compared with those predictors from questionnaire and biomarker assessment. I provide a framework for big data applications to obtain meaningful risk prediction and generate data-driven hypotheses.

## Data Availability

The data that support the findings of this study are available on request from the corresponding author, Xurui Jin. The data are not publicly available due to their containing information that could compromise the privacy of research participants.

## Notes

### Competing Interest Statement

The authors have declared no competing interest.

### Clinical Trial

N.A.

### Funding Statement

No external funding was received.

### Summary of Updates

Methods section revised

